# High Dietary Linoleic Acid Intake Suppresses Eicosapentaenoic Acid Status and Shifts Oxylipin Metabolism Towards Arachidonic Acid in Healthy Adults: A Randomized Controlled Trial

**DOI:** 10.64898/2026.04.09.26350499

**Authors:** Susan Sergeant, Linda Easter, Tammy Mustin, Priscilla Ivester, Jimaree Legins, Michael C. Seeds, Carrie S. Standage-Beier, Anderson Cox, Cristina M. Furdui, Brian Hallmark, Floyd H. Chilton

## Abstract

The modern Western diet (MWD) provides high linoleic acid (LA) exposure, typically contributing 6–9% of total caloric intake. These high LA levels have fueled a longstanding debate regarding whether this dietary pattern confers benefit or risk. Importantly, LA intake is disproportionately elevated among lower socioeconomic populations due to greater reliance on industrial seed oils and ultra-processed foods. Despite decades of research, controlled dietary intervention studies directly evaluating the biological consequences of varying LA exposure remain limited. The current randomized, double-blind intervention compared the effects of a 12-week Low LA diet (2.5% energy) versus a High LA diet (10.0% energy) in healthy adults. Primary outcomes included plasma highly unsaturated fatty acid (HUFA) concentrations and *ex vivo* zymosan-stimulated whole-blood oxylipin generation. Fifty-two participants completed the intervention. High LA exposure resulted in a marked reduction in plasma n-3 eicosapentaenoic acid (EPA) concentrations compared with the LowLA arm. In contrast, levels of arachidonic acid (ARA), dihomo-gamma-linolenic acid (DGLA) and docosahexaenoic acid (DHA) did not differ by dietary LA exposure. Analysis of oxylipin species revealed that levels of EPA-derived relative to ARA-derived mediators were significantly reduced in the High LA arm. These findings reveal that higher dietary LA selectively suppresses EPA pools and EPA-derived oxylipins without altering ARA, shifting the lipid mediator balance toward a more n-6-dominant profile.

## 1. Introduction

Over the past century, shifts in agricultural practices toward the industrial production of commodity crops, particularly soybeans and corn, have transformed the global food supply and launched a scientific and public health debate [1-4]. Central to this controversy is whether widespread dietary use of end products derived from these crops, including linoleic acid (LA)-rich vegetable oils is healthy. Elevation in dietary LA has contributed to a pervasive imbalance in dietary n-6 relative to n-3 polyunsaturated fatty acids (PUFA) that has been proposed to impact the incidence and progression of chronic diseases [5-7] This issue has expanded globally with the adoption of the Modern Western Diet (MWD), characterized by high intake of LA, which now constitutes a major component of both industrial food production and household cooking due to its low cost, long shelf life, and broad availability. Given their low cost, high presence in ultra-processed foods extensive use in fast-food outlets, and lack of access to healthy n-3-containing foods, the shift toward High LA commodity seed oils has disproportionately affected lower-income populations.

Early guidance for the increased use of dietary LA emerged from the diet–heart hypothesis of the 1960s–1970s, which proposed that replacing saturated fats with LA-rich vegetable oils would lower serum cholesterol, thereby reducing atherosclerosis and coronary heart disease risk. Consistent with this framework, early prospective randomized controlled trials and longitudinal dietary interventions were designed to increase LA intake, primarily through substitution of saturated fatty acids with vegetable oils [8-12]. However, initial reports from early randomized dietary trials were largely silent regarding the impact of LA-enriched interventions on other endpoints such as cardiovascular and all-cause mortality. A re-analysis of the Sydney Diet Heart Study showed that replacing saturated fat with high linoleic acid safflower oil markedly increased all-cause and cardiovascular mortality despite lowering cholesterol [13]. A related re-analysis of the Minnesota Coronary Experiment found no mortality benefit from lowering cholesterol via High LA corn oil, and a signal toward higher mortality in some subgroups [14]. And finally, a 2010 meta-analysis by Ramsden et al. distinguished mixed n-3/n-6 PUFA trials that showed reduced CHD risk from n-6–specific PUFA trials (primarily LA) which showed no benefit and possible harm [15]. Importantly, the meta-analysis did not claim definitive harm across all contexts but rather showed that benefits previously attributed to n-6 PUFA were largely driven by trials that increased n-3 alongside n-6.

Although typically present at lower dietary abundance, n-3 highly unsaturated fatty acids (HUFA), especially eicosapentaenoic acid (EPA) have been shown to play critical roles in regulating inflammation, thrombosis, and immune responses through its conversion into bioactive lipid mediators [16-18]. In contrast, metabolites of the n-6 HUFA, arachidonic acid (ARA), typically exhibit opposing biological activities, including promotion of inflammatory and pro- thrombotic signaling pathways [19,20]. Importantly, both EPA and ARA are synthesized by a common enzymatic pathway that includes precursor elongation, desaturation, membrane incorporation, and ultimately downstream HUFA oxylipin biosynthesis. Competition between n-6 and n-3 PUFAs along the elongation-desaturation pathway was first demonstrated in rat liver microsomal systems [21,22] and subsequent whole-animal feeding studies [23]. These studies confirmed that increasing dietary LA suppresses the conversion of alpha-linolenic acid (ALA) to EPA and docosahexaenoic acid (DHA) while reciprocally enhancing ARA synthesis from LA. Stable isotope tracer and controlled feeding studies in humans demonstrate that higher dietary LA reduces the conversion of ALA to long-chain n-3 HUFAs, particularly EPA [24-26].

Because LA and ALA compete for the same elongation and desaturation enzymes, variation in dietary LA intake has been shown in *in vitro* and *in vivo* animal models to shift the metabolic flux through this shared pathway, thereby altering membrane HUFA composition. Yet human data across physiologically relevant LA intake ranges remain limited. We therefore conducted a controlled clinical trial comparing High LA and Low LA dietary exposures to directly quantify how altering LA intake influences n-6 and n-3 HUFA synthesis, and the subsequent production of bioactive lipid mediators derived from EPA and ARA. This design enables an important assessment of how habitual LA intake modulates the biochemistry governing inflammatory and thrombotic signaling in humans.

## 2. Materials and Methods

This dietary intervention study was designed to examine the impact of the manipulation of dietary LA exposure on plasma PUFA and HUFA levels, and zymosan whole blood generated-oxylipin levels in healthy participants consuming a controlled diet containing either a Low (≤2.5% energy) or High (10% energy) LA dietary environment for 12 weeks (**Figure 1**). Naturally occurring variants of safflower, producing seed oils with low (∼13%) and high (∼75%) LA contents, were used to manipulate the dietary LA exposure. ALA exposure (∼1% energy), from flaxseed oil, was the consistent for both arms. The target calories from dietary fat exposure were 30% for both arms. A double-blinded, randomized parallel, two-arm dietary intervention study design was employed. Approval from Wake Forest University Health Sciences Institutional Review Board and the NIH/Office of Clinical Research Affairs was obtained before beginning the study. Th e trial was registered with ClinicalTrials.gov; NCT02962128.

**Figure 1:**
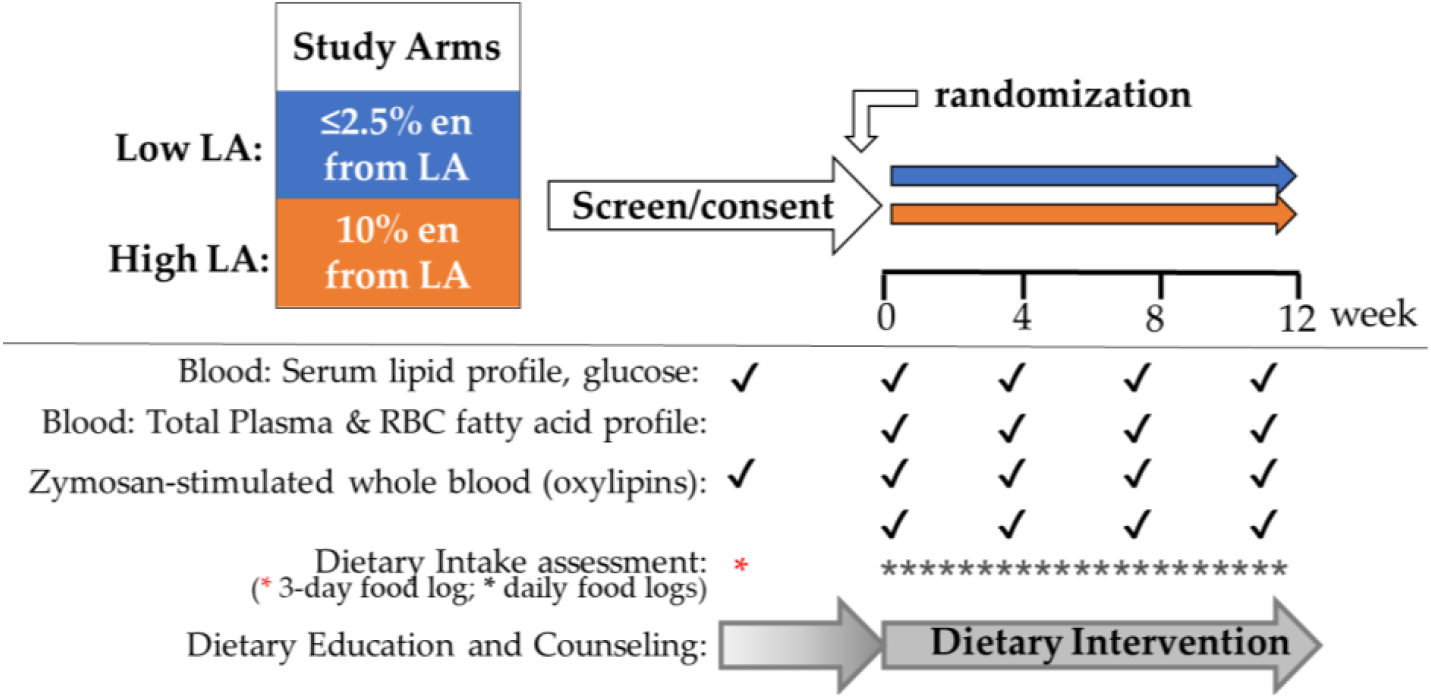
Study Design. The dietary intervention study was a double-blinded, randomized parallel-arm design. After consenting and a vigorous screening process, eligible participants were randomized one of two dietary interventions and consumed either a low LA (2.5% EN) or high LA (10% EN), with constant ALA exposure (2.5% EN) for both arms for 12 weeks. Participants met with Registered Dieticians (RD) weekly to pick up study foods, review weekly food logs and receive on-going dietary counselling. Participants met with the study team at 4-week intervals to provide fasting biospecimens and undergo a variety of anthropomorphic and cardiovascular measurements.

### 2.1 Study Participants

The study was conducted (August 2016 to December 2021) at the Wake Forest University Health Sciences (WFUHS). The inclusion criteria were healthy adults of European or African ancestry (self-identifying as non-Hispanic), 21-65 years of age and free of major diseases. Participant exclusion criteria included the following: a) current use of anti-inflammatory drugs (NSAIDs, oral/IV steroids, other injection anti-inflammatory drugs, >100mg aspirin per day, leukotriene receptor antagonists), niacin, fibrates or fish or botanical oils (containing PUFAs); b) blood pressure >140/90mm/Hg; c) fasting blood triglycerides >150mg/dL, fasting blood glucose > 125mg/dl; d) having a pacemaker, defibrillator or myocardial infarction, vascular surgery, or stroke in the preceding year; e) any stage heart failure, prior cholecystectomy, end stage renal disease; f) BMI <19 or >30; g), pregnancy or nursing; h) alcohol use >14 drinks per week; i) self-reported current use of tobacco, marijuana, or CBD use or other illicit drugs; and j) intolerance or allergy to safflower/flax oils or any component in provided study foods.

**Figure 2**. shows a summary of the recruitment and retention status for the study. For all participants, written informed consent was obtained during an initial screening visit. Since acceptable compliance to the dietary intervention had the potential to cause significant lifestyle changes, it was necessary for the study team and the registered dietitian (RD) to critically evaluate the suitably of each participant for the dietary challenges (adherence to dietary regime, regular eating habits, record keeping tasks, intervention duration, and limitations on travel during the intervention) before enrollment in the dietary intervention. Consented participants who met inclusion criteria and were determined, by the dietary staff, to be good candidates for this challenging dietary intervention were invited to join the intervention phase of the study. Block randomization was used to ensure approximately equal accrual to each sequence from each stratum over time. The Nutrition Coordinator (an RD; L. Easter) performed the randomization task using files provided by the study biostatistician to accomplish the block randomization. All other study staff and participants were blinded to dietary arm assignment.

### 2.2 Dietary Intervention

The study was designed to accurately capture PUFA dietary exposure [27] and to provide approximately 95% of daily fat calories. Prior to the beginning of the dietary intervention, participants underwent a fasting resting metabolic rate (RMR) test using a MGC Ultima CCM™ indirect calorimeter [28]. RDs also gathered information about typical eating habits (3-day food log), exercise habits, and food sensitivities. These data were evaluated to estimate the daily meal plan needed to meet the study nutrient goals and the caloric target needed to maintain body weight measured at that study visit. A weight change beyond ±5lbs of this weight during the intervention phase (based on weekly weights) required a change in the daily caloric intake value (typically in 200-calorie increments). Of the 80 participants beginning the dietary intervention, it was necessary to adjust the daily caloric goal for 20 participants, nearly equally distributed between the two study arms. Gathering information concerning eating habits and food sensitivities was imperative as it provided a means of identifying participants who would be poorly suited for participation from a dietary perspective. The study could not accommodate individuals with known allergies to study oils or gluten, on a vegan diet, lactose intolerant (unless already acclimated to lactase enzyme supplements), or any others whose lifestyle or preferences prevented compliance with the study guidelines.

**Figure 2:**
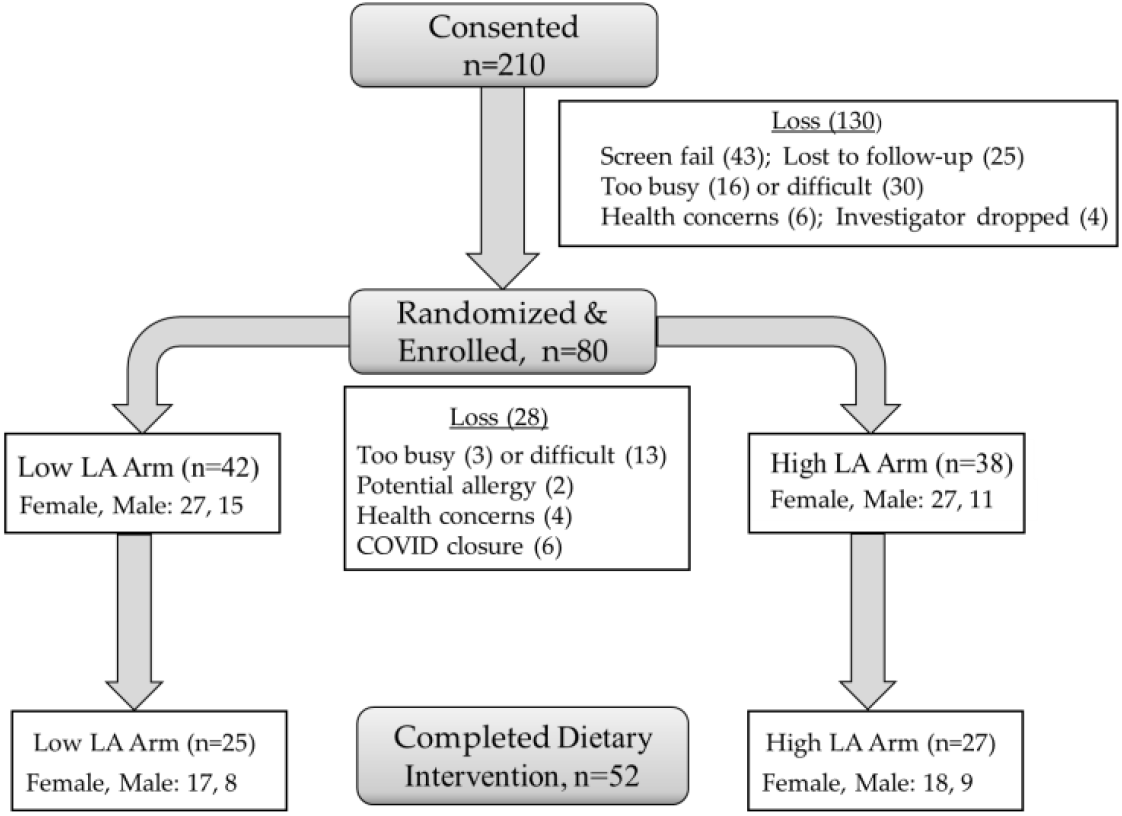
Recruitment and Retention. Participants were recruited from an independent Wake Forest University Health Sciences IRB-approved study seeking healthy adults, who agreed to be re-contacted for future studies and through local advertising campaigns. The nature of the 12-week dietary intervention (study provision of only a portion of daily foods and weekly study visits) was considered by most contacts to be too challenging since it often necessitated unwanted lifestyle changes. The counts for gender distribution are stated for those randomized (n=80) to and a total of 52 completing the entire 12-week intervention.

#### 2.2.1 Food Grade Oils

Oils constitute one of largest sources of dietary fat. Since the goal of the intervention was to manipulate the LA exposure of the two study arms, we took advantage of a pair of commonly available food-grade oils that differ in their LA content. Naturally occurring variants of safflower, having seed oils with low (∼13%) and high (∼73%) LA contents were used to achieve the manipulation of dietary LA exposure (**Supplemental Table S1**). In the low LA oil (high oleic acid safflower), oleic acid content of the safflower oils was much higher and the reciprocal of the LA content. Flaxseed oil was used as the source of ALA (∼56%) and was provided equally to both study arms.

Bulk oils were obtained from commercial suppliers after obtaining samples for analyses to evaluate their suitability (see below). A committed due diligence process was carried out to identify high quality food-grade oil products from trustworthy sources [29]. The oil sources were: Flaxseed oil (Shape Foods, Inc.; Brandon, Manitoba Canada); high oleic safflower oil (Oilseeds International Ltd.; San Francisco, CA, USA and Arista Industries Inc; Wilton, CT, USA); and high linoleic safflower oil (Oilseeds International Ltd.; San Francisco, CA, USA and Welch, Holme & Clark Co.; Newark, NJ, USA).

#### 2.2.2 Study-Provided Foods and Participant Educational Tools

During the dietary intervention, participants were provided (weekly) with a wide variety of study food items, prepared with the study oils containing the majority (>95%) of their daily dietary fat intake (25-30% of calories). All study-supplied foods were prepared, pre-weighed, packaged and provided by the WFUHS Clinical Research Unit (CRU) Metabolic Kitchen staff under research conditions. Daily smoothies (3-4 flavor recipes offered) were used as a vehicle for flaxseed oil, which was common to both arms. This delivery mechanism avoided heat-induced oxidation to this oil. Other provided study foods included study oil-based condiments (salad dressings, mayonnaise, and sauces), and a variety of prepared snack foods (cookie, brownie, granola, sweet/salty snack mixes, hummus), each made with the arm-specific type of safflower oil. A can of PAM™ aerosolized vegetable oil was dispensed, as needed, for participants to use in cooking. The recipes of all provided food items were designed to contribute a standard number of calories to the daily diet.

Participants ordered their preferred study food items on a weekly basis. All other allowed foods, including low/non-fat animal products (with controlled amounts of specific seafoods), grains, fruits, vegetables, fat-free sweets, and non-caloric beverages, were provided by the participants. An individualized meal plan was designed for each participant with the combined input of the RD and the participant. The input of the latter was essential to best accommodate their typical eating pattern and dietary preferences within the study nutrient requirements. The meal plans provided participants with a guide for planning all meals, which in turn facilitated their compliance and adherence to this challenging intervention protocol. It was critical for participants to avoid additional fat calories from other food types (dairy, meats, baked goods, etc.) to remain compliant with the study protocol. No run-in period on a common diet was employed for this study. Instead, the study RD and her team provided extensive, arm-independent, one-on-one dietary education and counseling as an essential element of the study protocol. RDs delivered a variety of assistance including individualized meal-planning assistance, allowed/disallowed food lists, grocery-shopping guides, instructions for interpreting food labels for fat content determination, guidance for home preparation of meals, guidance for meal selection when dining out, and estimation of portion sizes. Intensive RD-delivered counselling was provided prior to beginning the dietary intervention and continued weekly thereafter during food pick-up visits. Other education, tools, and counselling interactions provided to participants by the RDs included: a) lists of allowed and disallowed foods, b) food label reading practice (to select allowed foods and avoid extra fat exposure from grocery store products), c) use of a food scale (study provided) to measure portions, d) sample menu plans, e) recipes and suggestions to enhance food item acceptability, f) guidance concerning dining out to remain compliant, g) safe food handling procedures, and h) guided modification of the individualized meal plan, as needed. All dietary information was delivered by the RDs in a uniform manner to all participants.

### 2.3 Biospecimen, Collection, Handling and Storage

At the initial screening/consenting visit, blood was collected to obtain serum for screening exclusion criteria (lipid panel, glucose, hs-CRP). The remaining serum was aliquoted and stored at -70°C for future use. At subsequent visits, a dedicated tube of heparinized blood was collected at each fasting visit during the dietary intervention (weeks 0, 4, 8, and 12 of the intervention). Within 30 min of collection, the whole blood was subjected to a zymosan-stimulation assay for the evaluation of oxylipin generation [30-32]. Briefly, whole blood was incubated at 37°C for 30min in the presence of phosphate buffered saline or zymosan (Sigma-Aldrich, St. Louis, MO, USA) suspended in the same buffer (2.5mg/ml, final concentration). At the end of the incubation period, plasma was collected, aliquoted, blanketed with argon and stored at -70°C for oxylipin mass spectrometry analysis (below).

### 2.4 Measurements

During the dietary intervention, vital signs (blood pressure and resting heart rate) and morphometric measurements (waist and hip circumference, height; body mass index (BMI) and percent body fat) were obtained at 4-week intervals. Participants were weighed weekly by the RD staff at food pickup visits. The primary outcomes were circulating levels of n-6 and n-3 PUFAs and HUFA, ratios of circulating n-6 and n-3 HUFAs and levels and ratios of ARA- and EPA-derived lipoxygenase-derived oxylipins.

#### 2.4.1 Compliance

Compliance to the intervention was monitored in two ways. Dietary exposure to LA was obtained from weigh-backs of returned, uneaten study foods and from food logs completed by participants in real time and returned to the RD at weekly visits. At these visits, the RD queried the participant about any incomplete entries to obtain sufficient detail for calculating nutrient intake. The logs were analyzed using Nutrition Data System for Research (NDSR) developed at the University of Minnesota Nutrition Coordinating Center [33,34]. Cumulative nutrient intake data used to evaluate compliance included: percentage of calories as LA, ALA, total fat, carbohydrates and protein. The second measure of compliance was circulating fatty acid profiles measured from fasting blood samples obtained at 4-week intervals during the intervention.

#### 2.4.2 Fatty Acid Analysis in Plasma, Oils and Study Foods

Total plasma fatty acids were analyzed from fasting blood. Fatty acid including LA, ALA, oleic acid (OA), gamma-linoleinc acid (GLA), dihomo-gamma-linolenic acid (DGLA), ARA, stearidonic acid (SDA), EPA docosapentaenoic acid, (DPA), DHA were analyzed as fatty acid methyl esters (FAME). FAMEs were prepared [35] after alkaline hydrolysis of complex lipids in duplicate samples (100μl of plasma) in the presence of an internal standard (triheptade-canoin: NuChek Prep, Elysian, MN, USA; included for purposes of fatty acid quantification) as previously described [36,37]. A standard panel of 25 fatty acids (which accounted for 99% of the fatty acids in the samples) was quantified by gas chromatography with flame ionization detection (GC-FID) using an Agilent 7890 instrument with DB-23 column (30m, 0·25 mm ID, 0·25 µm film) fitted with an inert pre-column (1 m, 0.53 mm ID) for cool on-column injection. Data capture and analyses were performed with ChromPerfect software (v6.0.12). The instrument response factor has been calculated based on the use of external standard sets for quality assurance purposes and a mixture of known FAMEs was run with each sample set to monitor instrument performance [38]. Individual fatty acids in plasma were expressed as concentrations.

The fatty acid composition of the oil supplements (**Supplemental Table S1**) was determined in aliquots of oil diluted (in hexane) and processed as described above in the presence of the internal standard. For the purposes of product authentication, individual fatty acids in oil products were expressed as area% (**Supplemental Table S1**) to facilitate comparison to vendor documentation of product fatty acid profile.

The integrity of the study oils was monitored over time by routine evaluation of fatty acid content and oxidation status to ensure that a high-quality product was provided to the Metabolic Kitchen for study food preparation. For the oxidation status of oils, standard food industry assays were used. These included: a) peroxide value (PV; primary oxidants; iodometric titration assay [39]; European Pharmacopeia 2.5.5); b) anisidine value (AV; secondary oxidants; colorimetric assay [40]; European Pharmacopeia 2.5.36), and Totox (total oxidation = 2PV+AV). Oil was considered out of spec when the Totox value was ≥ 30. Such oil containers were removed from use and discarded.

The fatty acid profiles of the study food items were also monitored qualitatively over the course of the study as a quality control tool to ensure that the correct oils were used for the intended arm. A finite weight of food product (50-150mg, depending on oil content of the food item) was processed as described above for FAME analysis in the absence of the internal standard. It was not possible to perform oxidation assay on study food items due to the presence of interfering substances in these samples.

#### 2.4.3 Oxylipin Analysis

Oxylipins were extracted from plasma (100µl) in methanol (LC-MS grade) in the presence of deuterated internal standards. Briefly, the methanolic extracts were diluted in 9 volumes of water, centrifuged at room temperature to remove precipitated proteins. The supernatant was subjected to solid phase extraction (Phenomenex; Strata X-33µm Polymeric Reverse Phase; #8B-S100-EBJ) using a vacuum manifold. The column was pre-conditioned (1ml methanol followed by 1ml water) before loading the sample supernatant. After the sample had slowly eluted from the column, the column was washed once with 10% methanol. Oxylipins were eluted with 1ml of methanol and the eluant was collected in 1.5ml screw cap tubes and blanketed with argon and stored at -80ºC. Samples were prepared for analysis by drying under a stream of nitrogen, redissolving in 50 µl water: methanol (1:1).

Separation of the analytes was performed with a 150 × 3mm × 2.6 um Phenomenex Kinetex C8 column (PN: 00F-4497-Y0). The liquid chromatography system consisted of Shimadzu (Kyoto, Japan) LC-40D x3 solvent delivery modules, a DGU-405 degasser, an SCL-40 system controller, and a CTO-40C column oven. Analytes were eluted with a mobile phase system consisting of Fisher OmniSolv water with 0.1% formic acid (Fisher) as solvent A and 100% Fisher OmniSolv acetonitrile for solvent B. Solvents flow was set to 0.4 ml/min and a starting point of 30% B. This gradient was increased to 95% B over 9 minutes and then held until 11 minutes. The concentration was then decreased back to 30% B at 11.1 minutes and held until the method ended. The mass spectrometer, an AB SCIEX 7500 triple quadrupole (SciEx, Framingham, MA), was run in negative ion mode utilizing nitrogen gas for ion generation under the following conditions: Ion source gas 1 and 2 were set at 60 and 90 psi, respectively, the curtain gas was set at 40psi and the CAD gas was set to 12. The source temperature was set to 350 °C. Ion spray voltage was set to 2000 V.

#### 2.4.4 Clinical Analysis

A dedicated tube of blood for the preparation of serum was collected at each fasting study visit (the initial screening/consenting visit and each of the four monthly visits during the dietary intervention). Routine cardiometabolic and inflammatory biomarkers were measured as secondary outcomes. These included: 1) serum derived from fasting blood was used to measure glucose (enzymatic assay), 2) high sensitivity C-reactive protein (hsCRP; immunochemoluminometric assay), and 3) serum lipids (total cholesterol, triglycerides, HDL-, VLDL- and LDL-cholesterol; enzymatic assays). These endpoints were analyzed by a qualified clinical laboratory (Lab Corp, Burlington, NC).

### 2.5 Data Analysis

Statistical analyses were conducted in GraphPad (version 5). Additional analyses were performed in STATA (version 11.1) for mixed-effect modeling to evaluate potential arm and time (study week) interactions. Fatty acid data was evaluated as concentration (mg/dl), but log transformed as needed. The zymosan-stimulated oxylipin generation was calculated as the abundance (pg/ml) in the +zymosan less that in the buffer control condition. The resultant zymosan-stimulated oxylipin data was cleaned of missing or negative values. Oxylipins having <15% missing data were used for further analyses. Statistical comparisons were made by 2-tailed t-test at each time point by study arm. Analyses were not adjusted for age or sex as covariates.

## 3. Results

### 3.1 Characteristics of the study population

The target study population consisted of healthy adults. As illustrated in **Figure 2**, 80 consented participants met eligibility criteria and were enrolled in the dietary intervention phase. Following randomization, 16 participants withdrew, primarily due to scheduling constraints or difficulty adhering to the intervention protocol. Notably, participants who reported challenges with maintaining mindful daily dietary tracking were more likely to demonstrate poor compliance. Additional attrition occurred due to suspected study-related food allergy (n=2), emerging health concerns unrelated to the intervention (n=4), and an institutional pause of research activities during the COVID-19 pandemic (n=6). Ultimately, 52 participants completed the full 12-week dietary intervention: 25 in the Low-LA arm and 27 in the High LA arm.

Baseline demographic and clinical characteristics of the 80 randomized participants are presented in **Table 1**. The two intervention arms were comparable with respect to initial sample size, sex distribution, and racial composition. Among age, anthropometric measures, cardiovascular indices, and standard clinical laboratory parameters, only fasting HDL cholesterol and fasting glucose differed modestly between groups at baseline.

**Table 1.**
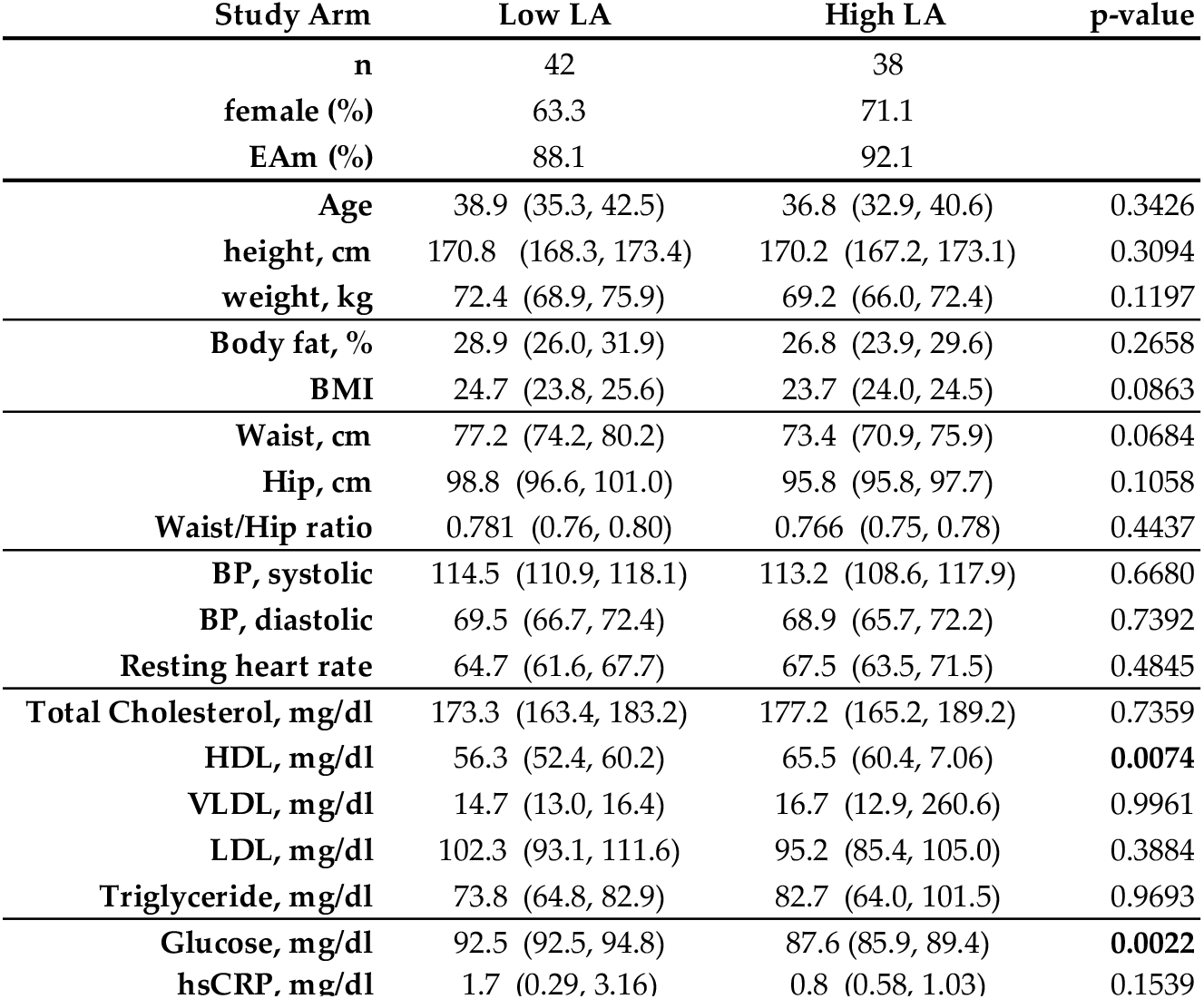
Baseline Demographics. The demographics of the study cohort (n=80) at baseline (week 0 of dietary intervention) are shown by dietary intervention arm. The data are mean (with 95% confidence intervals). Data were analyzed by 2-tailed T-test and significant arm differences at baseline are indicated by a bolded p-value.

| Study Arm | Low LA | High LA | p-value |
| --- | --- | --- | --- |
| **n** | 42 | 38 |  |
| **female (%)** | 63.3 | 71.1 |  |
| **EAm (%)** | 88.1 | 92.1 |  |
| **Age** | 38.9 (35.3, 42.5) | 36.8 (32.9, 40.6) | 0.3426 |
| **height, cm** | 170.8 (168.3, 173.4) | 170.2 (167.2, 173.1) | 0.3094 |
| **weight, kg** | 72.4 (68.9, 75.9) | 69.2 (66.0, 72.4) | 0.1197 |
| **Body fat, %** | 28.9 (26.0, 31.9) | 26.8 (23.9, 29.6) | 0.2658 |
| **BMI** | 24.7 (23.8, 25.6) | 23.7 (24.0, 24.5) | 0.0863 |
| **Waist, cm** | 77.2 (74.2, 80.2) | 73.4 (70.9, 75.9) | 0.0684 |
| **Hip, cm** | 98.8 (96.6, 101.0) | 95.8 (95.8, 97.7) | 0.1058 |
| **Waist/Hip ratio** | 0.781 (0.76, 0.80) | 0.766 (0.75, 0.78) | 0.4437 |
| **BP, systolic** | 114.5 (110.9, 118.1) | 113.2 (108.6, 117.9) | 0.6680 |
| **BP, diastolic** | 69.5 (66.7, 72.4) | 68.9 (65.7, 72.2) | 0.7392 |
| **Resting heart rate** | 64.7 (61.6, 67.7) | 67.5 (63.5, 71.5) | 0.4845 |
| **Total Cholesterol, mg/dl** | 173.3 (163.4, 183.2) | 177.2 (165.2, 189.2) | 0.7359 |
| **HDL, mg/dl** | 56.3 (52.4, 60.2) | 65.5 (60.4, 70.6) | **0.0074** |
| **VLDL, mg/dl** | 14.7 (13.0, 16.4) | 16.7 (12.9, 20.6) | 0.9961 |
| **LDL, mg/dl** | 102.3 (93.1, 111.6) | 95.2 (85.4, 105.0) | 0.3884 |
| **Triglyceride, mg/dl** | 73.8 (64.8, 82.9) | 82.7 (64.0, 101.5) | 0.9693 |
| **Glucose, mg/dl** | 92.5 (92.5, 94.8) | 87.6 (85.9, 89.4) | **0.0022** |
| **hsCRP, mg/dl** | 1.7 (0.29, 3.16) | 0.8 (0.58, 1.03) | 0.1539 |

Importantly, baseline (Day 0) fasting plasma n-6 and n-3 PUFA and HUFA concentrations (mg/dL) were similarly distributed between the two study arms (**Supplemental Table S2**). Furthermore, plasma PUFA-to-HUFA ratios used as surrogate markers of enzymatic activity within the shared elongation and desaturation biosynthetic pathway did not differ between groups at baseline, supporting metabolic equivalence prior to dietary intervention.

### 3.2 Impact of the dietary intervention

The objective of manipulating dietary LA exposure to determine PUFA-to-HUFA metabolism in healthy, free-living adults posed a substantial design challenge, as typical U.S. dietary intake of LA ranges from approximately 6–9% of total energy intake. Accordingly, a dietary intervention was engineered to sustain markedly divergent LA exposures with a goal of 10% of total energy in the High LA arm and 2.5% of total energy in the Low LA arm.

The two dietary exposure targets were successfully achieved and maintained over the 12-week intervention period (**Figure 3**). LA intake during the intervention was 2.65 ± 0.06% energy in the Low LA arm and 10.21 ± 0.09% energy in the High LA arm (**Figure 3a**). Importantly, pre-intervention (baseline) LA intake did not differ between study arms (**Figure 3a, open symbols**). Following initiation of the intervention (**Figure 3a, closed symbols**), LA exposure was significantly reduced in the Low LA group and significantly increased in the High LA group, confirming successful execution of the intended dietary manipulation.

**Figure 3:**
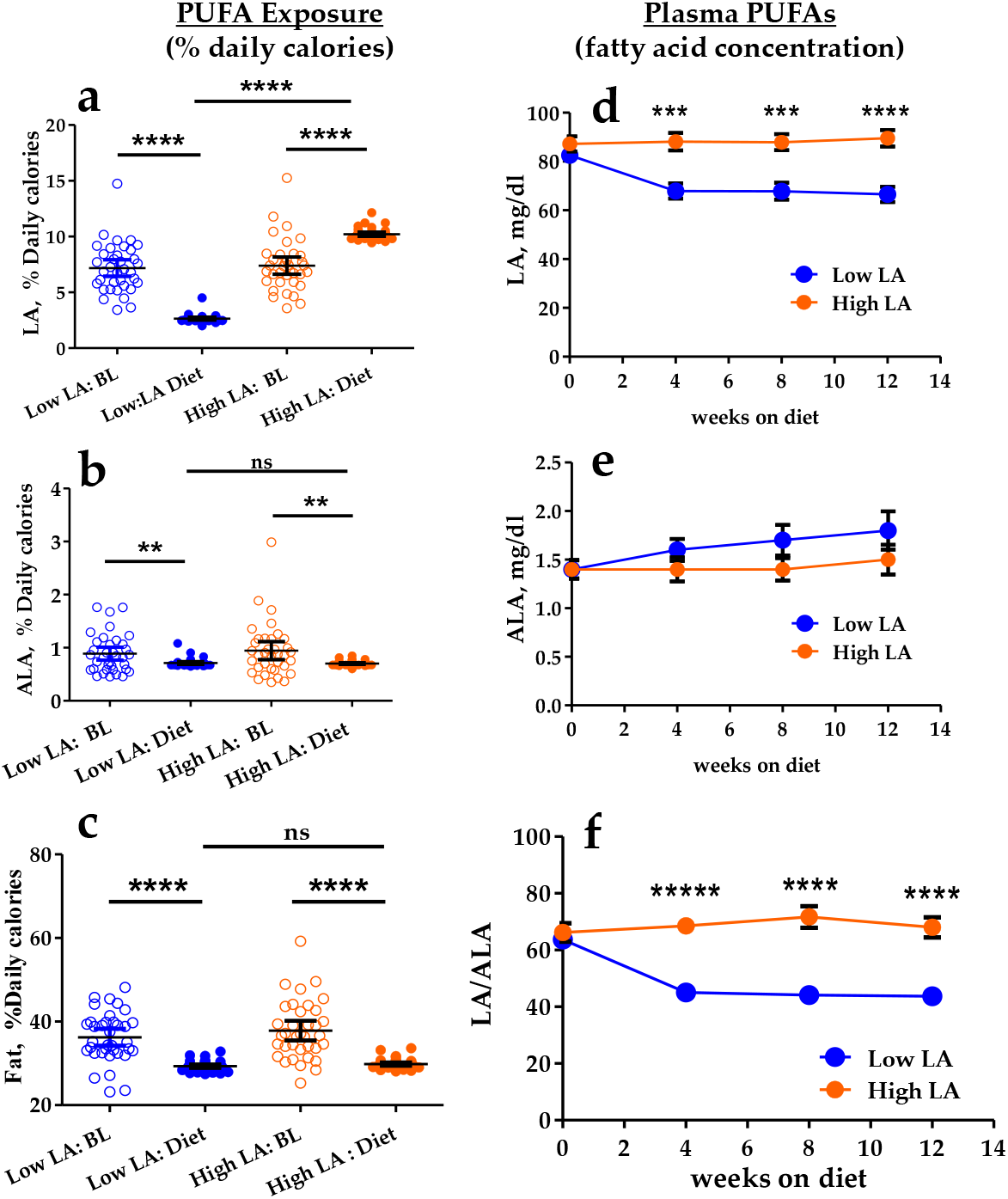
Effects of Dietary Intervention on LA Exposure and Plasma PUFA Concentrations. Dietary intakes of LA (a), ALA (b), and total fat (c) expressed as percentage of daily caloric intake. Pre-intervention baseline (BL) estimates were derived from three-day food logs collected prior to the intervention (open symbols). Compliance during the intervention was assessed from weekly diet logs (closed symbols). Data represent means ± 95% CI; group differences were assessed by two-tailed Student’s t-test. Fasting plasma concentrations (mg/dL) of LA (d), ALA (e), and the LA/ALA ratio (f) over the course of the dietary intervention are shown for both study arms. Data represent means ± SEM. Between-group comparisons at each time point were made by two-tailed Student’s t-test. Significance levels: **, p < 0.01; ***, p < 0.001; ****, p < 0.0001; *****, p < 1×10^-7^

In contrast, the ALA exposure (<2.5% EN) remained constant for both dietary arms (**Figure 3b; closed symbols**). Notably, the total fat caloric intake was not quantitatively different between the two dietary arms during the intervention.

However, both arms showed an approximate 20% decrease in fat calorie consumption during the dietary intervention compared to baseline consumption (**Figure 3c; closed symbols**). There was no commensurate arm-dependent difference in body weight, body fat or BMI by the end of the intervention since the total daily calories were not different between arms. The diet design only imposed quantitative changes in LA exposure and an attendant reciprocal change in oleic acid (OA) exposure due to the high oleic acid safflower oil (data not shown).

Dietary compliance was further validated using fasting plasma PUFA concentrations (mg/dL). As shown in **Figure 3d**, the elevated dietary LA exposure in the High LA arm resulted in significantly elevated circulating plasma LA concentrations compared with the Low LA arm at weeks 4, 8, and 12 (t-test, p = 1× 10−^4^ to 7.25 × 10−^6^; χ^2^ for Arm-study week interaction, p<0.00001). In contrast, the controlled ALA intake produced no between-arm differences in fasting plasma ALA concentrations throughout the intervention (**Figure 3e**; χ^2^ for Arm-study week interaction, p=0.4592). Consequently, the plasma LA/ALA ratio diverged between arms, with the High LA group demonstrating a significantly greater ratio by week 4 that was sustained through week 12 (**Figure 3f**). Importantly, the circulating LA/ALA ratio closely mirrored dietary exposure ratios derived from food record compliance data (High LA, 14.5; Low LA, 3.7). For context, the average pre-intervention LA/ALA ratio was 7.9, consistent with typical Modern Western dietary patterns.

### 3.3 Impact of LA Exposure on PUFA to HUFA Biosynthesis

As outlined in the Introduction, the PUFA-to-HUFA biosynthetic pathway proceeds through alternating desaturation and elongation steps catalyzed sequentially by FADS2 (Δ6-desaturase), ELOVL5 (elongase5), and FADS1 (Δ5-desaturase). Because n-6 and n-3 PUFAs share this enzymatic machinery, the pathway represents a critical node of metabolic competition between the two fatty acid families. A significant arm-dependent divergence was observed at both the ELOVL5 and FADS1 steps of the n-3 branch, resulting in markedly lower plasma concentrations of the n-3 HUFAs eicosatetraenoic acid (ETA) and EPA in the High LA arm (**Figure 4**). This suppression of n-3 HUFA production is consistent with elevated LA competitively displacing n-3 substrates at key enzymatic steps. Although not reaching statistical significance, plasma concentrations of DPA and DHA also trended lower in the High LA group, suggesting that the suppressive effect of elevated LA may extend further along the n-3 elongation pathway.

**Figure 4:**
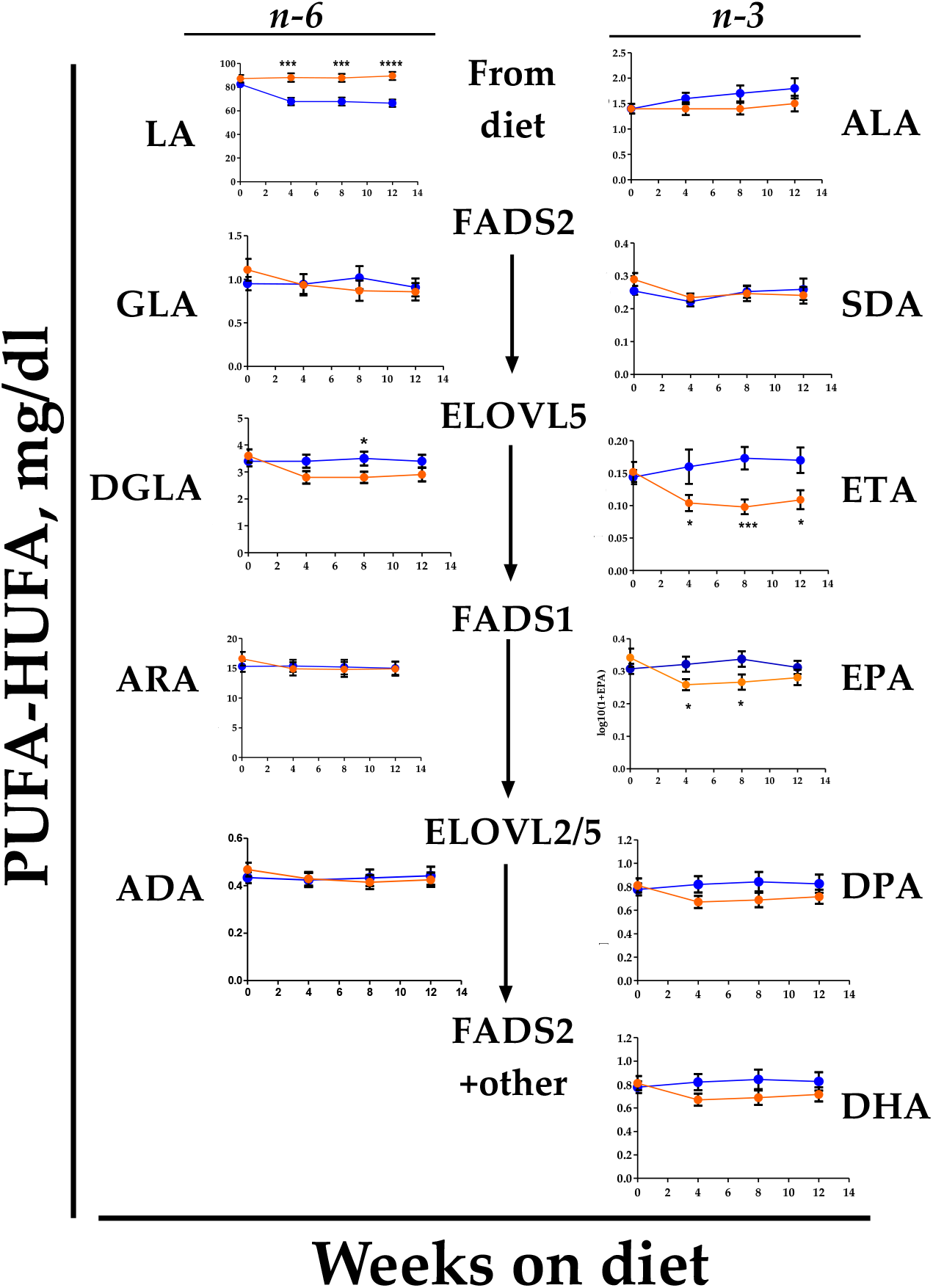
Plasma PUFA and HUFA Concentration Across Their Biosynthetic Pathway. Fasting concentrations (as mg/dl) for paired n-6 and n-3 PUFA or HUFA at each pathway enzymatic step are shown. Adrenic acid (ADA). Data are presented as mean±sem data and statistical comparisons were made by 2-tailed T-test at each time point by study arm. P-values are represented as: *, p<0.05 **, p<0.01; ***, p<1×10^-3^; ****, p <1×10^-4^.

In contrast, the n-6 HUFAs DGLA and ARA did not differ between arms (**Figure 4**), indicating that approximately 2.5% of energy from LA is sufficient to sustain near-maximal flux through the n-6 branch of the pathway. The absence of any further increase in DGLA or ARA at 10% energy LA reinforces this interpretation, suggesting a saturation of n-6 HUFA biosynthesis at the lower intake level. An arm-study week interaction was highly significant for ETA (chi2, p=0.0068), the EPA precursor.

Regression analyses of EPA (**Figure 5a**), DHA (**Figure 5b**), and DGLA concentrations (**Figure 5c**) as a function of ARA concentrations across study weeks 4–12 revealed a markedly LA-dependent relationship for EPA (F = 27.85; p < 0.0001), but not for DHA (F = 4.48; p = 0.036) or DGLA (F = 0.084, p=0.773. This analysis further supported the conclusion that elevated dietary LA selectively and potently suppresses EPA accumulation relative to ARA.

**Figure 5:**
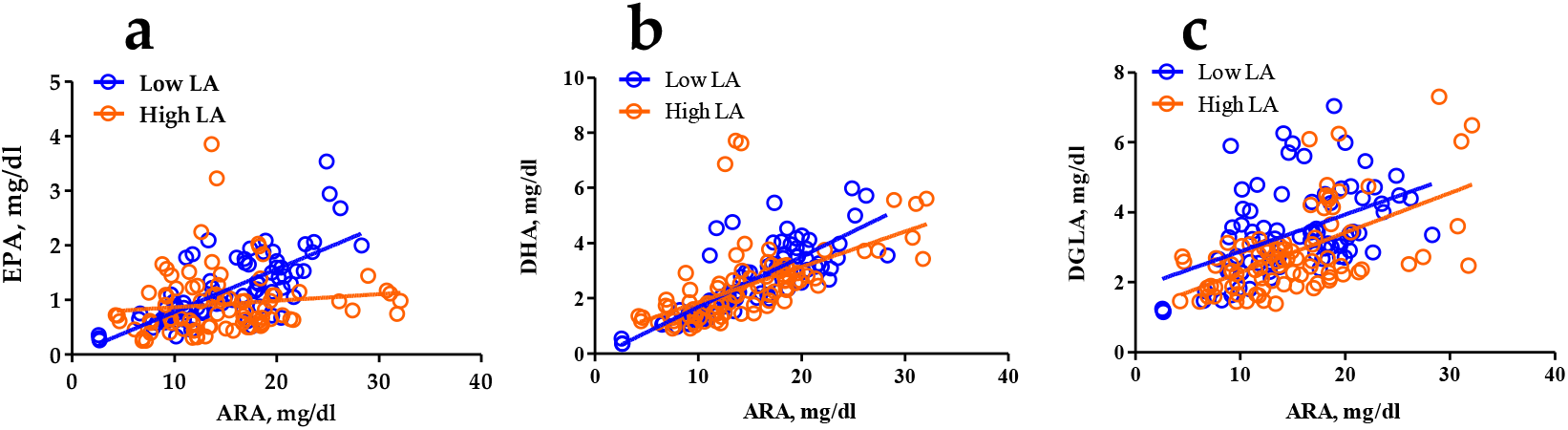
Plasma EPA Concentrations Are Arm-Dependent. A regression analyses between plasma EPA and ARA (**a**), DHA and ARA (**b**) or DGLA and ARA (**c**) concentrations (mg/dl) for intervention weeks (4,8,12) are shown by study arm. The slopes of the regression lines for EPA were significantly different: EPA-ARA, (F=27.85, p<0.0001) but much less so for DHA-ARA, (F=4.48 p=0.036) and not significant for DGLA (F=0.084, p=0.773).

Consistent with the marked reduction in EPA relative to ARA, the plasma ARA/EPA ratio was significantly elevated in the High LA arm (**Figure 6a**; (χ^2^, p=0.0006), further supporting the conclusion that n-3 HUFA biosynthesis is selectively suppressed under conditions of elevated n-6 substrate competition. The specificity of the n-3 effect is underscored by the absence of significant differences in the ARA to DGLA ratios between arms (**Figures 6b and 6c**).

**Figure 6:**
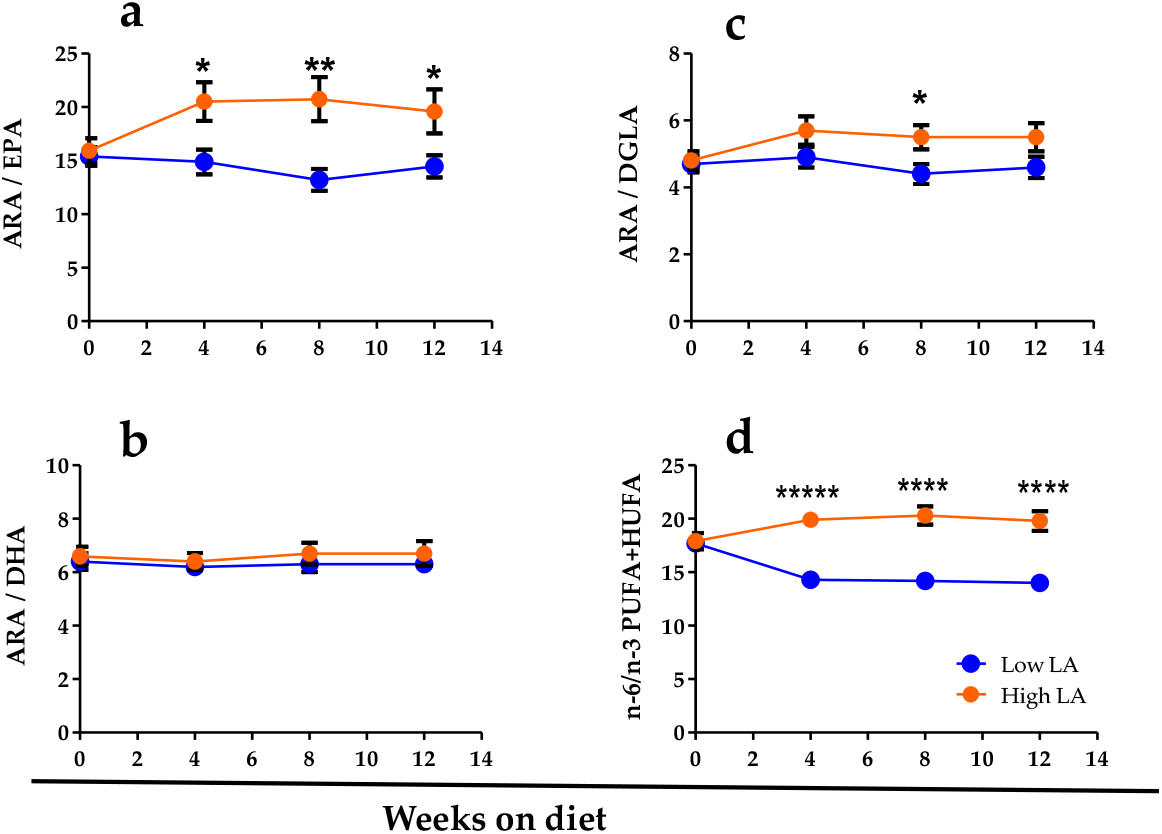
Impact of Dietary LA on HUFA Ratios. HUFA ratio between ARA and EPA(**a**), DHA (**b**) and DGLA (**c**) or total n-6/n-3 (**d**) were calculated from fatty acid concentrations (mg/dl). Data are shown as mean±sem. Statistical comparisons were made by 2-tailed t-test at each study time point. Significant differences by t-test. P-values are represented as follows: *, p<0.05; **, p<0.01; ***, < p < 1×10^-3^; ****, p < 1×10^-4^; *****, p < 1×10^-7^.

Importantly, the arm-driven shift in n-3 HUFA synthesis was consistent over time as shown as total n-6 to n-3 PUFA+HUFA ratio (**Figure 6d**; (χ^2^, p<1.00001). This ratio was markedly higher in the High LA arm, reflecting both the elevated n-6 substrate load and the concomitant suppression of n-3 HUFA production along the shared biosynthetic pathway. This broad shift in the n-6 to n-3 balance highlighted that the metabolic consequences of high dietary LA extend beyond any single fatty acid.

### 3.4 Impact of dietary LA in oxylipin generation

Most assessments of oxylipins in human disease have focused on circulating species measured in plasma or serum, where oxylipins exist in both non-esterified (free) and esterified (phospholipid and lipoprotein-bound) pools [41-43]. However, given the large number of enzymatic (LOX, COX, CYP) and non-enzymatic pathways contributing to oxylipin generation, their rapid turnover, metabolism, and dynamic interconversion between esterified and free pools, it remains uncertain how well static circulating concentrations reflect pathway flux or functional inflammatory potential.

To address this limitation, the current study evaluated the impact of dietary LA exposure on oxylipin production capacity using a zymosan-stimulated whole blood assay, thereby capturing integrated leukocyte-driven enzymatic responses under a standardized inflammatory challenge, with a particular emphasis on lipoxygenase (LOX)-derived products. Targeted LC–MS/MS profiling yielded high-quality quantitative data for 40 oxylipin species, which were included in subsequent analyses. Among oxylipins generated in the zymosan-stimulated assay, 35% (14 species) were derived from ARA, 25% (10 species) from EPA, 15% (6 species) from LA, 10% each from DGLA and DHA, and one species each from ALA and GLA (**Figure 7a**).

**Figure 7:**
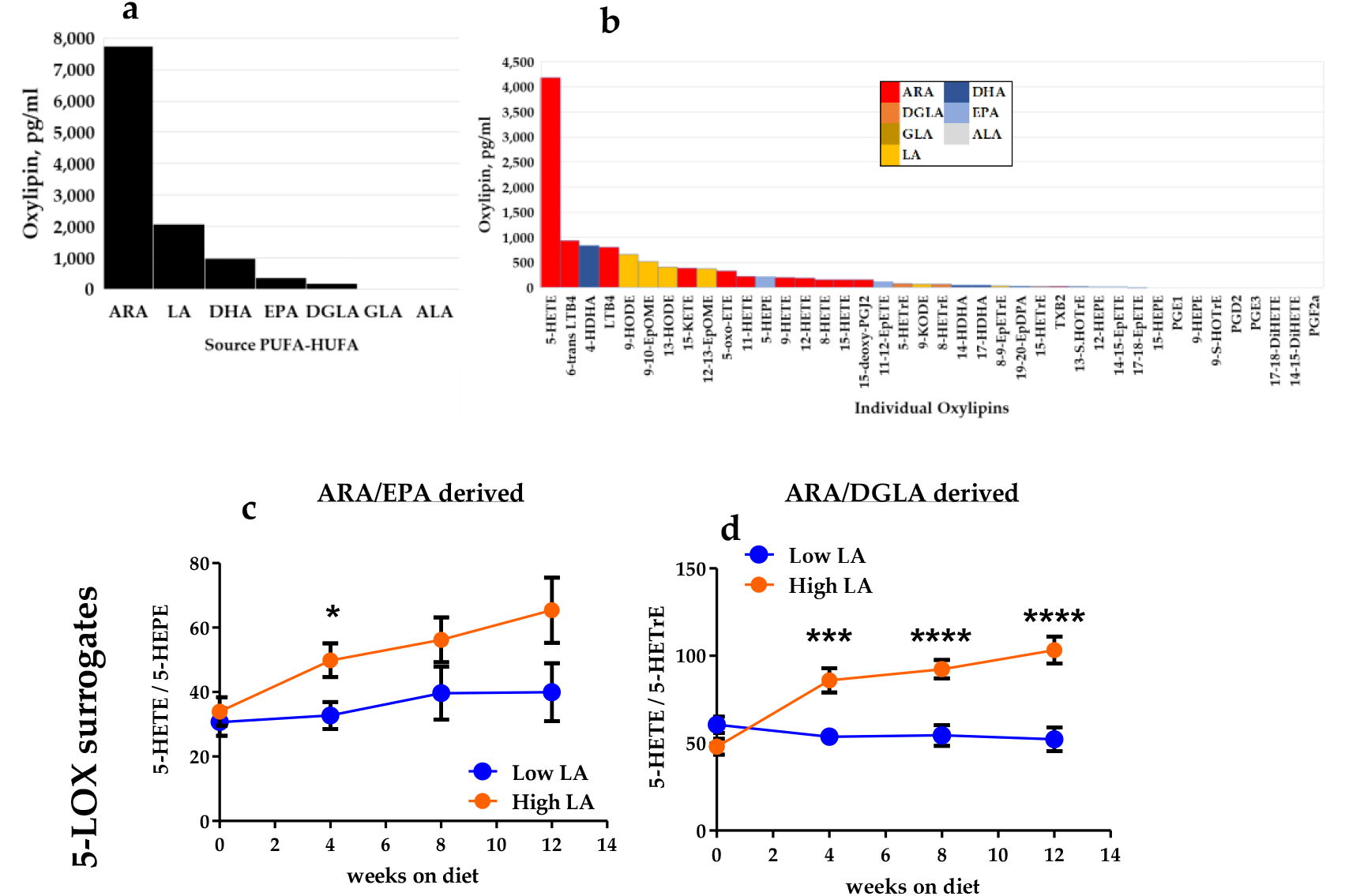
Abundance of Oxylipins Generated by Zymosan-Stimulation of Whole Blood. Oxylipins extracted from plasma derived from zymosan-stimulated whole blood of those participants (n=52) were analyzed by targeted LC-MS/MS. The abundance of oxylipins (pg/ml) by source PUFA or HUFA were ranked (**a**). The abundance of individual oxylipins (pg/ml plasma; mean value) generated from zymosan-stimulated whole blood is shown (**b**). The oxylipin ratio of ARA-derived 5-HETE to EPA-derived 5 HEPE are shown in **(c)** and ARA-derived 5-HETE to DGLA-derived 5-HETrEare shown in (**d**). Significant differences by p-value are represented as follows: *, p<0.05; **, p<0.01; ***, <p < 1×10^-3^; ****, p < 1×10^-4^.

Quantitative analyses of individual species confirmed predominance of a 5-lipoxygenase (5-LOX)-mediated response, with marked enrichment of 5-hydroxyeicosatetraenoic acid (5-HETE) among ARA-derived products (**Figure 7b**). Among EPA-derived metabolites, 5-hydroxy-eicosapentaenoic acid (5-HEPE) was the most abundant species. Low abundance oxylipins (<50 pg/mL) are presented in **Supplemental Figure S1**. 5-LOX also utilizes DGLA and its 5-LOX product is 5-hydroxy-6E,8Z,11Z-eicosatrienoic acid, HETrE). The 5-HETE/5-HEPE ratio (ARA/EPA) and 5-HETE/5-HETrE ratio (ARA/DGLA) reflect the utilization n-3 versus n-6 HUFA precursors. Both ratios were consistently higher in the High LA arm (**Figures 7c 7d**), indicating significantly elevated 5-LOX production from ARA compared to both EPA (χ^2^, p=0.0124) and DGLA (χ^2^, p <0.00001).

## 4. Discussion

This controlled dietary intervention demonstrates that LA, within ranges commonly consumed in modern diets, functions as a dominant regulator of the human PUFA-to-HUFA metabolic network. Increasing LA from ∼2.5% to ∼10% of total energy selectively suppressed endogenous n-3 HUFA synthesis, reduced circulating EPA concentrations, increased the ARA/EPA ratio, and shifted leukocyte 5-LOX metabolism toward preferential ARA-derived 5-HETE formation under an inflammatory challenge. These findings provide direct human evidence that high LA intake does not merely contribute to membrane composition but establishes competitive control over a shared enzymatic pathway with downstream consequences for inflammatory signaling capacity.

### 4.1 Major Challenge of this Study

The n-6 and n-3 PUFA families share a finite sequence of desaturation and elongation enzymes. Although substrate competition within this pathway has been inferred from *in vitro* and animal studies [21,22], human data across physiologically relevant LA exposures have remained sparse. A major challenge in conducting such studies is the dietary control required to achieve meaningfully distinct LA exposure levels, particularly the low end of the range. The 2.5% energy LA target achieved in the Low LA arm of this study represents a substantially reduced LA intake compared to typical Western diets and required considerable dietary intervention to sustain.

The logistical demands to achieve this level of dietary control were considerable. Rigorous compliance monitoring and intensive participant support were necessary throughout the intervention period to maintain adherence to the prescribed diets, and the study required substantial staffing resources to achieve this. Despite these efforts, 16% of participants withdrew from the study, predominantly from the Low LA arm (data not shown), reflecting the challenge participants faced in needing to make lifestyle changes in order to adhere to substantially modified dietary patterns during the intervention period. The need to maintain adequate staff-to-participant ratios to ensure dietary compliance necessarily constrained the overall sample size of the study to n=80.

### 4.2 LA as a Competitive Metabolic Gatekeeper

Here, ∼2.5% energy from LA was sufficient to sustain circulating ARA and DGLA levels, suggesting that near-maximal n-6 HUFA synthesis is achieved at relatively modest LA intake. In contrast, elevating LA to ∼10% energy selectively suppressed the n-3 branch, with significant reductions in total plasma concentrations of ETA and EPA and a consistent downward trend in DPA, while DHA remained unchanged. This is agreement with earlier studies that challenged humans with LA exposure and evaluated EPA in plasma lipid fractions [44,45] and the established enzymatic loci of n-6 and n-3 PUFA to HUFA competition [46].

This asymmetric response reveals a hierarchical architecture: additional LA does not further increase n-6 HUFA production but instead redistributes flux away from n-3 HUFA synthesis. Within the intake range studied, LA therefore behaves less as a simple nutrient and more as a competitive gatekeeper of pathway allocation. Importantly, these data suggest that levels of LA intake found within the MWD may operate well beyond the threshold required for n-6 sufficiency and instead also function to constrain endogenous n-3 HUFA production.

### 4.3 Functional Recalibration of Inflammatory Signaling

The shifts in HUFA levels were paralleled by functional changes in oxylipin generation capacity. Using a zymosan-stimulated whole-blood assay, we captured integrated leukocyte enzymatic responses under standardized activation conditions. The dominance of 5-LOX–derived products reflect the central role of this pathway in acute inflammatory signaling. Because 5-LOX utilizes ARA, EPA, and DGLA as substrates, product ratios directly index enzymatic substrate competition. Elevated LA intake increased both the 5-HETE/5-HEPE (ARA/EPA) and 5-HETE/5-HETrE (ARA/DGLA) ratios, indicating preferential metabolism of ARA under high LA exposure. The impact of the high LA diet in the 5-HETE/5HETrE ratio was not expected and may represent a compartmentalization of 5-LOX substrates that is being impacted by high LA levels.

### 4.4 Rethinking “Total PUFA”

ALA intake and total PUFA exposure were held constant between arms; yet, marked divergence emerged in HUFA composition and oxylipin output. These results challenge the prevailing practice of grouping PUFAs as a single functional class [47]. These data reveal that total PUFA quantity does not predict metabolic outcome; rather, subclass competition determines pathway flux. Within the intake range examined, increasing LA did not enhance n-6 HUFA abundance but disproportionately suppressed n-3 HUFA generation, thereby altering the n-6/n-3 balance. This competitive topology implies that metabolic effects of LA cannot be inferred from cholesterol reduction or total PUFA metrics alone. Network redistribution effects must be considered.

### 4.5 EPA as the Metabolically Sensitive Node

The selective suppression of EPA, with preservation of DHA, identifies EPA as the most diet-responsive node within the endogenous n-3 pathway under competitive LA exposure. DHA synthesis or concentrations requires additional elongation, desaturation, and peroxisomal processing steps [48]. DHA concentrations may also be more highly influenced by DHA exposure, which may buffer against substrate competition. Alternatively, DHA pools may be preferentially conserved due to structural demands. The dissociation between EPA and DHA responses suggests that endogenous EPA may be particularly vulnerable to high LA exposure, with potential implications for interpreting circulating EPA as a biomarker of dietary fatty acid balance and inflammatory capacity.

### 4.6 Context Within the Ongoing LA Debate

The role of dietary LA in cardiometabolic health remains debated. Observational and interventional studies have frequently reported inverse associations between LA intake and LDL cholesterol or cardiovascular risk, leading to recommendations favoring replacement of saturated fat with n-6 PUFAs [8-12]. However, such analyses rarely account for competitive interactions within the shared PUFA-to-HUFA pathway or for downstream redistribution of n-3 substrate flux. The present findings do not address clinical outcomes directly; rather, they identify a mechanistic effect of LA on metabolic network allocation. Within physiologically relevant intake ranges, increasing LA does not further augment ARA abundance but instead selectively constrains endogenous n-3 HUFA synthesis and shifts oxylipin generation toward ARA-derived products. These results suggest that metabolic consequences of LA exposure cannot be fully inferred from selected clinical metrics alone and underscore the importance of considering competitive pathway topology when interpreting dietary PUFA data.

These mechanistic considerations carry particular relevance for socioeconomically disadvantaged populations. Low-income communities in the United States face disproportionate reliance on ultra-processed foods which are predominantly formulated with high-LA seed oils as a function of cost, availability, and food environment constraints. Food insecurity and SNAP participation are both associated with higher consumption of ultra-processed foods, with adults experiencing very low food security consuming the highest proportions [49]. Simultaneously, participants with lower educational attainment and income had significantly lower n-3 PUFA and HUFA intakes and fish intakes across NHANES 2003–2014 survey cycles [50]. This is a consistent pattern suggesting that socioeconomically disadvantaged populations are at heightened risk for lower n-3 HUFA intake, which may further contribute to health disparities. This convergence of high LA exposure and low n-3 PUFA and HUFA intake is the dietary environment in which the competitive pathway suppression of EPA synthesis demonstrated here would be expected to be most consequential.

### 4.7 Strengths and Limitations

This study integrates controlled dietary manipulation, objective biochemical compliance validation, quantitative lipidomics, and functional oxylipin profiling under standardized immune activation. The experimental isolation of LA as the primary manipulated variable strengthens causal inference. Limitations include moderate sample size, 12-week duration, reliance on circulating lipid pools, and stimulated rather than *in vivo* inflammatory assessments. Whether sustained LA-driven flux redistribution influences long-term disease trajectories remains to be determined.

## 5. Conclusions

Within physiologically relevant intake ranges, dietary LA exerts competitive control over the shared PUFA desaturation pathway, selectively suppressing endogenous EPA synthesis and biasing leukocyte 5-LOX metabolism toward ARA-derived mediators. These findings demonstrate that dietary fatty acid composition can reallocate metabolic flux and recalibrate inflammatory signaling capacity through substrate competition within a common enzymatic network.

## Data Availability

All data produced in the present study are available upon reasonable request to the authors unless the study has been closed at the level of the Institutional Review Board.

## Supplementary Materials

Table S1: Fatty Acid Profile of Study Oils;

Table S2: Baseline Plasma PUFA-HUFA Profile by Arm

Figure S1: Low Abundance Oxylipins Generated by Zymosan-Stimulated in Whole Blood

## Author Contributions

S.S.: investigation, methodology, supervision, formal analysis, visualization, writing—original draft preparation, review and editing; L.H.E.: methodology, investigation; T.M.: investigation, methodology; P.I.: investigation, methodology; J.L.: formal analysis; M.C.S.: data curation, validation; C.S-B.: visualization; A.O.C.: methodology; C.M.F.: funding acquisition; B.H.: formal analysis; F.H.C.: Conceptualization, funding acquisition, writing—review and editing, supervision. All authors have read and agreed to the published version of the manuscript. All authors have read and agreed to the published version of the manuscript.

## Institutional Review Board Statement

The study was conducted in accordance with the Declaration of Helsinki and reviewed and approved by the Institutional Review Board of The Wake Forest University Health Sciences (IRB00038046, approved 8/18/16) and by the NIH/Office of Clinical Research Affairs. (Clinicaltrial.gov, NCT02962128).

## Informed Consent Statement

Informed consent was obtained from all subjects involved in the study.

## Data Availability Statement

The de-identified data presented in this study are available upon request to the corresponding author unless the study has been closed at the level of the Institutional Review Board.

## Acknowledgments

We greatly appreciate and acknowledge the spirit of all the study participants who tackled this most challenging of dietary interventions. Registered dietitians Mary Self and Bryce Alford provided continued and encouraging dietary counseling to assist participants in their adherence to the intervention protocol throughout the study. The authors acknowledge the support of the Atrium Health Wake Forest Baptist Proteomics and Metabolomics Shared Resource, supported by the National Cancer Institute’s Cancer Center Support Grant award number P30CA012197.

## References

1. Mukhopadhyay, R. An essential debate: A controversy over a dietary recommendation for omega-6 fatty acids shows no sign of resolving itself. ASBMB Today 2012, 11, 14–19.

2. Gibson, R.A. Musings about the role dietary fats after 40 years of fatty acid research. Prostaglandins Leukot Essent Fatty Acids 2018, 131, 1–5, doi:10.1016/j.plefa.2018.01.003.

3. Lands, B. Consequences of Essential Fatty Acids. Nutrients 2012, 4, 1338–1357, doi:10.3390/nu4091338.

4. Marklund, M.; Wu, J.H.Y.; Imamura, F.; Del Gobbo, L.C.; Fretts, A.; de Goede, J.; Shi, P.; Tintle, N.; Wennberg, M.; Aslibekyan, S., et al. Biomarkers of Dietary Omega-6 Fatty Acids and Incident Cardiovascular Disease and Mortality. Circulation 2019, 139, 2422–2436, doi:10.1161/circulationaha.118.038908.

5. Blasbalg, T.L.; Hibbeln, J.R.; Ramsden, C.E.; Majchrzak, S.F.; Rawlings, R.R. Changes in consumption of omega-3 and omega-6 fatty acids in the United States during the 20th century. Am J Clin Nutr 2011, 93, 950–962, doi:doi:10.3945/ajcn.110.006643.

6. Simopoulos, A.P. The importance of the omega-6/omega-3 fatty acid ratio in cardiovascular disease and other chronic diseases. Exp Biol Med 2008, 233, 674–688, doi:doi:10.3181/0711-MR-311.

7. Simopoulos, A.P. The importance of the ratio of omega-6/omega-3 essential fatty acids. Biomed Pharmacother 2002, 56, 365–379, doi:10.1016/s0753-3322(02)00253-6.

8. Woodhill, J.M.; Palmer, A.J.; Leelarthaepin, B.; McGilchrist, C.; Blacket, R.B. Low fat, low cholesterol diet in secondary prevention of coronary heart disease. Adv Exp Med Biol 1978, 109, 317–330, doi:10.1007/978-1-4684-0967-3_18.

9. Frantz, I.D., Jr.; Dawson, E.A.; Ashman, P.L.; Gatewood, L.C.; Bartsch, G.E.; Kuba, K.; Brewer, E.R. Test of effect of lipid lowering by diet on cardiovascular risk. The Minnesota Coronary Survey. Arteriosclerosis 1989, 9, 129–135, doi:10.1161/01.atv.9.1.129.

10. Rose, G.A.; Thomson, W.B.; Williams, R.T. Corn Oil in Treatment of Ischaemic Heart Disease. Br Med J 1965, 1, 1531–1533, doi:10.1136/bmj.1.5449.1531.

11. Turpeinen, O.; Karvonen, M.J.; Pekkarinen, M.; Miettinen, M.; Elosuo, R.; Paavilainen, E. Dietary prevention of coronary heart disease: the Finnish Mental Hospital Study. Int J Epidemiol 1979, 8, 99– 118, doi:10.1093/ije/8.2.99.

12. JN, M.; etal. Controlled Trial of Soya-bean Oil in Myocardial Infarction: Report of a Research Committee to the Medical Research Council. The Lancet 1968, 292, 693–700, doi:10.1016/S0140-6736(68)90746-0.

13. Ramsden, C.E.; Zamora, D.; Leelarthaepin, B.; Majchrzak-Hong, S.F.; Faurot, K.R.; Suchindran, C.M.; Ringe, l.; Davis, J.M.; Hibbeln, J.R. Use of dietary linoleic acid for secondary prevention of coronary heart disease and death: evaluation of recovered data from the Sydney Diet Heart Study and updated meta-analysis. BMJ 2013, 346, doi:10.1136/bmj.e8707, doi:10.1136/bmj.e8707.

14. Ramsden, C.E.; Zamora, D.; Majchrzak-Hong, S.; Faurot, K.R.; Broste, S.K.; Frantz, R.P.; Davis, J.M.; Ringel, A.; Suchindran, C.M.; Hibbeln, J.R. Reevaluation of the traditional diet-heart hypothesis: analysis of recovered data from Minnesota Coronary Experiment (1968-73). BMJ 2016, 353, i1246, doi:10.1136/bmj.i1246.

15. Ramsden, C.E.; Hibbeln, J.R.; Majchrzak, S.F.; Davis, J.M. n-6 Fatty acid-specific and mixed polyunsaturate dietary interventions have different effects on CHD risk: a meta-analysis of randomised controlled trials. Br J Nutr 2010, 104, 1586–1600.

16. Calder, P.C. Omega-3 fatty acids and inflammatory processes: from molecules to man. Biochem Soc Trans 2017, 45, 1105–1115, doi:10.1042/BST20160474.

17. Serhan, C.N. Pro-resolving lipid mediators are leads for resolution physiology. Nature 2014, 510, 92– 101, doi:10.1038/nature13479.

18. Bhatt, D.L.; Steg, P.G.; Miller, M.; Brinton, E.A.; Jacobson, T.A.; Ketchum, S.B.; Doyle, R.T.; Juliano, R.A.; Jiao, L.; Granowitz, C., et al. Cardiovascular Risk Reduction with Icosapent Ethyl for Hypertriglyceridemia. N Engl J Med 2019, 380, 11–22, doi:10.1056/NEJMoa1812792.

19. Funk, C.D. Prostaglandins and leukotrienes: advances in eicosanoid biology. Science 2001, 294, 1871– 1875, doi:10.1126/science.294.5548.1871.

20. Chilton, F.H.; Murphy, R.C.; Wilson, B.A.; Sergeant, S.; Ainsworth, H.C.; Seeds, M.C.; Mathias, R.A. Diet-gene interactions and PUFA metabolism: A potential contributor to health disparities and human diseases. Nutrients 2014, 6, 1993–2022, doi:doi:10.3390/nu6051993.

21. Lands, W.E.; Morris, A.; Libelt, B. Quantitative effects of dietary polyunsaturated fats on the composition of fatty acids in rat tissues. Lipids 1990, 25, 505–516, doi:10.1007/BF02537156.

22. Sprecher, H.; Luthria, D.L.; Mohammed, B.S.; Baykousheva, S.P. Reevaluation of the pathways for the biosynthesis of polyunsaturated fatty acids. J Lipid Res 1995, 36, 2471–2477.

23. Leece, E.A.; Allman, M.A. The relationships between dietary alpha-linolenic:linoleic acid and rat platelet eicosapentaenoic and arachidonic acids. Br J Nutr 1996, 76, 447–452, doi:10.1079/bjn19960049.

24. Emken, E.A.; Adlof, R.O.; Gulley, R.M. Dietary linoleic acid influences desaturation and acylation of deuterium-labeled linoleic and linolenic acids in young adult males. Biochim Biophys Acta - Lipids Lipid Metab 1994, 1213, 277–288, doi:10.1016/0005-2760(94)00054-9.

25. Burdge, G.C.; Wootton, S.A. Conversion of alpha-linolenic acid to eicosapentaenoic, docosapentaenoic and docosahexaenoic acids in young women. Br J Nutr 2002, 88, 411–420, doi:10.1079/BJN2002689.

26. Hussein, N.; Ah-Sing, E.; Wilkinson, P.; Leach, C.; Griffin, B.A.; Millward, D.J. Long-chain conversion of [13C]linoleic acid and a-linolenic acid in response to marked changes in their dietary intake in men. J.Lipid Res. 2005, 46, 269–280.

27. MacIntosh, B.A.; Ramsden, C.E.; Faurot, K.R.; Zamora, D.; Mangan, M.; Hibbeln, J.R.; Mann, J.D. Low-n-6 and low-n-6 plus high-n-3 diets for use in clinical research. Br J Nutr 2013, 111, 559–568, doi:10.1017/S0007114512005181.

28. Black, C.; Grocott, M.P.; Singer, M. Metabolic monitoring in the intensive care unit: a comparison of the Medgraphics Ultima, Deltatrac II, and Douglas bag collection methods. Br J Anaesth 2015, 114, 261–268, doi:10.1093/bja/aeu365.

29. Sorkin, B.C.; Kuszak, A.J.; Williamson, J.S.; Hopp, D.C.; Betz, J.M. The Challenge of Reproducibility and Accuracy in Nutrition Research: Resources and Pitfalls. Advances in Nutrition 2016, 7, 383–389, doi:10.3945/an.115.010595.

30. Fradin, A.; Zirrolli, J.A.; Maclouf, J.; Vausbinder, L.; Henson, P.M.; Murphy, R.C. Platelet-activating factor and leukotriene biosynthesis in whole blood. A model for the study of transcellular arachidonate metabolism. Journal of Immunology 1989, 143, 3680–3685.

31. Wheelan, P.; Murphy, R.C. Quantitation of 5-lipoxygenase products by electrospray mass spectrometry: effect of ethanol on zymosan-stimulated production of 5-lipoxygenase products by human neutrophils. Analytical Biochemistry 1997, 244, 110–115, doi:10.1006/abio.1996.9885.

32. Hester, A.G.; Murphy, R.C.; Uhlson, C.; Ivester, P.; Lee, T.C.; Sergeant, S.; Miller, L.R.; Howard, T.D.; Mathias, R.A.; Chilton, F.H. Relationship between a common variant in the fatty acid desaturase (FADS) cluster and eicosanoid generation in humans. J Biol Chem 2014, 289, 22482–22489, doi:10.1074/jbc.M114.579557.

33. Feskanich, D.; Sielaff, B.H.; Chong, K.; Buzzard, I.M. Computerized collection and analysis of dietary intake information. Comput Methods Programs Biomed 1989, 30, 47–57, doi:10.1016/0169-2607(89)90122-3.

34. Sievert, Y.A.; Schakel, S.F.; Buzzard, I.M. Maintenance of a nutrient database for clinical trials. Control Clin Trials 1989, 10, 416–425, doi:10.1016/0197-2456(89)90006-8.

35. Metcalfe, L.D.; Schmitz, A.A.; Pelka, J.R. Rapid preparation of fatty acid esters from lipids for gas chromatographic analysis. Anal Chem 1966, 38, 514–515.

36. Sergeant, S.; Hallmark, B.; Mathias, R.A.; Mustin, T.L.; Ivester, P.; Bohannon, M.L.; Johnstone, L.; Seeds, M.; Chilton, F.H. Prospective clinical trial examining the impact of genetic variation in FADS1 on the metabolism of linoleic acid- and gamma-linolenic acid-containing botanical oils. Am J Clin Nutr 2020, nqaa023, doi:doi:10.1093/ajcn/nqaa023.

37. Weaver, K.L.; Ivester, P.; Seeds, M.C.; Case, L.D.; Arm, J.; Chilton, F.H. Effect of dietary fatty acids on inflammatory gene expression in healthy humans. J Biol Chem 2009, 284, 15400–15407.

38. Brenna, J.T.; Plourde, M.; Stark, K.D.; Jones, P.J.; Lin, Y.-H. Best practices for the design, laboratory analysis, and reporting of trials involving fatty acids. Am J Clin Nutr 2018, 108, 211–227, doi:10.1093/ajcn/nqy089 %J The American Journal of Clinical Nutrition.

39. Europe, C.o. European Pharmacopoeia Commission Directorate for the Quality of M. 2.5 Assays. In European Pharmacopoeia, 7th Edition ed.; Healthcare, D.f.t.Q.o.M., Ed. Council of Europe: Strasbourg, 2010; pp. 137–138.

40. Talbot, G. 16 - The Stability and Shelf Life of Fats and Oils. In The Stability and Shelf Life of Food (Second Edition), Subramaniam, P., Ed. Woodhead Publishing: 2016; pp. 461–503.

41. Shearer, G.C.; Walker, R.E. An overview of the biologic effects of omega-6 oxylipins in humans. Prostaglandins, Leukotrienes and Essential Fatty Acids 2018, 137, 26–38, doi:10.1016/j.plefa.2018.06.005.

42. Quehenberger, O.; Armando, A.M.; Brown, A.H.; Milne, S.B.; Myers, D.S.; Merrill, A.H.; Bandyopadhyay, S.; Jones, K.N.; Kelly, S.; Shaner, R.L., et al. Lipidomics reveals a remarkable diversity of lipids in human plasma. J Lipid Res 2010, 51, 3299–3305, doi:10.1194/jlr.M009449.

43. Psychogios, N.; Hau, D.D.; Peng, J.; Guo, A.C.; Mandal, R.; Bouatra, S.; Sinelnikov, I.; Krishnamurthy, R.; Eisner, R.; Gautam, B., et al. The Human Serum Metabolome. PLOS ONE 2011, 6, e16957, doi:10.1371/journal.pone.0016957.

44. Liou, Y.; King, D.; Zibrik, D.; Innis, S. Decreasing Linoleic Acid with Constant α-Linolenic Acid in Dietary Fats Increases (n-3) Eicosapentaenoic Acid in Plasma Phospholipids in Healthy Men. J Nutr 2007, 137, 945–952.

45. Taha, A.Y.; Cheon, Y.; Faurot, K.F.; MacIntosh, B.; Majchrzak-Hong, S.F.; Mann, J.D.; Hibbeln, J.R.; Ringel, A.; Ramsden, C.E. Dietary omega-6 fatty acid lowering increases bioavailability of omega-3 polyunsaturated fatty acids in human plasma lipid pools. Prostaglandin Leukotr Essential Fatty Acid 2014, In Press, doi:10.1016/j.plefa.2014.02.003.

46. Lands, B.; Bibus, D.; Stark, K.D. Dynamic interactions of n-3 and n-6 fatty acid nutrients.Prostaglandins Leukot Essent Fatty Acids 2018, 136, 15–21, doi:10.1016/j.plefa.2017.01.012.

47. Ramsden, C.E.; Hibbeln, J.R.; Majchrzak-Hong, S.F. All PUFAs are not created equal: absence of CHD benefit specific to linoleic acid in randomized controlled trials and prospective observational cohorts. World Rev Nutr Diet 2011, 102, 30–43, doi:10.1159/000327789.

48. Sprecher, H. Metabolism of highly unsaturated n-3 and n-6 fatty acids. Biochim Biophy Acta - Mol Cell Biol Lipids 2000, 1486, 219–231, doi:10.1016/S1388-1981(00)00077-9.

49. Leung, C.W.; Fulay, A.P.; Parnarouskis, L.; Martinez-Steele, E.; Gearhardt, A.N.; Wolfson, J.A. Food insecurity and ultra-processed food consumption: the modifying role of participation in the Supplemental Nutrition Assistance Program (SNAP). Am J Clin Nutr 2022, 116, 197–205, doi:10.1093/ajcn/nqac049.

50. Cave, C.; Hein, N.; Smith, L.M.; Anderson-Berry, A.; Richter, C.K.; Bisselou, K.S.; Appiah, A.K.; Kris-Etherton, P.; Skulas-Ray, A.C.; Thompson, M., et al. Omega-3 Long-Chain Polyunsaturated Fatty Acids Intake by Ethnicity, Income, and Education Level in the United States: NHANES 2003-2014. Nutrients 2020, 12, doi:10.3390/nu12072045.

