## Supplemental Data for "High Dietary Linoleic Acid Intake Suppresses Eicosapentaenoic Acid Status and Shifts Oxylipin Metabolism Towards Arachidonic Acid in Healthy Adults: A Randomized Controlled Trial"

**Supplemental Figure S1**: Low **Abundance Oxylipins Generated by Zymosan-Stimulation of Whole Blood**

This figure shows an expanded portion of the right side of **Figure 7b** to highlight the lower abundance (>50pg/ml) oxylipins generated during the whole blood stimulation of immune cells by the phagocytic stimulus, zymosan.

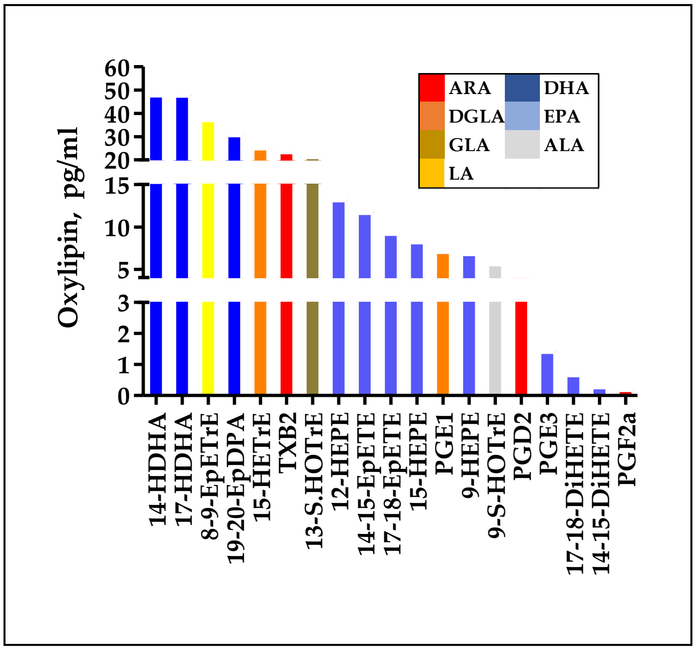

**Supplemental Table S1: Fatty Acid Profile of Dietary Study Oils.** The fatty acid composition of the naturally occurring safflower oils were used to manipulate LA exposure. Flax seed oil served as the source of ALA. The data is presented as area%. The measured fatty acids account for >99% of those in the oils. Bolded fatty acids indicated those targeted by the dietary manipulation.

| | | **Study Oil FA Profile (area%)** | | |
|---|---|---|---|---|
| | | *Safflower* | | |
| **Common Name** | **FA** | **Low LA** | **High LA** | **Flaxseed** |
| Myristolate | C14:0 | <0.1 | <0.1 | <0.1 |
| Palmitic | C16:0 | 5.4 | 7.2 | 5.5 |
| Palmitoleic | C16:1 | 0 | 0 | <0.1 |
| Stearic | C18:0 | 2.7 | 3.5 | 4.2 |
| **Oleic (OA)** | C18:1 n-9c | **77.4** | **14.5** | **18.8** |
| Vaccenic | C18:1 n-7c | 0.5 | 0.5 | 0.5 |
| **Linoleic (LA)** | C18:2w n-6 | **12.8** | **73.1** | **14.5** |
| $\gamma$-Linolenic | C18:3 n-6 | 0.2 | 0.2 | 0.1 |
| **$\alpha$-Linolenic (ALA)** | C18:3 n-3 | **0.2** | **0.3** | **56.2** |
| Stearidonic | C18:4 n-3 | <0.1 | 0 | <0.1 |
| Arachidic | C20:0 | 0.3 | 0.1 | <0.1 |
| Gondoic | C20:1 n-9 | 0.2 | <0.1 | <0.1 |
| Behenic | C22:0 | 0.1 | 0.1 | <0.1 |
| Erucic | C22:1 n-9 | 0 | <0.1 | 0.0 |
| Nervonic | C24:1 n-9 | 0.1 | 0.1 | 0.0 |
| Others | | <0.1 | <0.1 | <0.1 |
| **Total** | | 99.9 | 99.8 | 99.9 |

**Supplemental Table S2: Baseline Plasma n-6 and n-3 PUFA and HUFA Concentrations in Each Arm.** Baseline (Week 0 of intervention) plasma PUFA, HUFA and MUFA concentrations (mg/dl) by study arm and ratios are shown. Data are presented as mean values with the 95% confidence interval. Data from the study arms were analyzed by 2-tailed T-test with the resultant p-value stated.

| | Mean | 95% CI | *p-value* | | Mean | 95% CI | *p-value* |
|---|---|---|---|---|---|---|---|
| ***n-6 PUFA-HUFA*** | | | | ***n-3 PUFA-HUFA*** | | | |
| **LA** | | | | **ALA** | | | |
| **Low LA** | 82.5 | 77.4, 87.6 | 0.232 | **Low LA** | 1.421 | 1.2, 1.6 | 0.833 |
| **High LA** | 87.2 | 19.1, 93.5 | | **High LA** | 1.448 | 1.2, 1.6 | |
| **GLA** | | | | **EPA** | | | |
| **Low LA** | 0.951 | 0.80, 1.1 | 0.274 | **Low LA** | 1.09 | 0.9, 1.3 | 0.938 |
| **High LA** | 1.11 | 0.9, 1.4 | | **High LA** | 1.1 | 0.9, 1.3 | |
| **DGLA** | | | | **DPA** | | | |
| **Low LA** | 3.42 | 3.1, 4.1 | 0.541 | **Low LA** | 0.78 | 0.7, 0.9 | 0.668 |
| **High LA** | 3.609 | 3.1, 4.1 | | **High LA** | 0.81 | 0.7, 0.9 | |
| **ARA** | | | | **DHA** | | | |
| **Low LA** | 15.3 | 13.5, 17.2 | 0.459 | **Low LA** | 2.63 | 2.2, 3.0 | 0.509 |
| **High LA** | 16.32 | 14.2, 18.4 | | **High LA** | 2.87 | 2.2, 3.5 | |
| ***n-6 PUFA-HUFA Ratios*** | | | | ***n-3 PUFA-HUFA Ratios*** | | | |
| **GLA/LA** | | | | **EPA/ALA** | | | |
| **Low LA** | 0.011 | 0.01, 0.013 | 0.453 | **Low LA** | 0.83 | 0.69, 0.97 | 0.951 |
| **High LA** | 0.012 | 0.01, 0.015 | | **High LA** | 0.84 | 0.65, 1.02 | |
| **DGLA/GLA** | | | | **DHA/ALA** | | | |
| **Low LA** | 4.78 | 3.8, 5.7 | 0.441 | **Low LA** | 1.96 | 1.65, 2.27 | 0.761 |
| **High LA** | 4.29 | 3.5, 5.1 | | **High LA** | 2.03 | 1.65, 2.41 | |
| **ARA/DGLA** | | | | **DHA/EPA** | | | |
| **Low LA** | 4.68 | 4.2, 5.2 | 0.841 | **Low LA** | 2.55 | 2.22, 2.88 | 0.6 |
| **High LA** | 4.75 | 4.2, 5.3 | | **High LA** | 2.68 | 2.30, 3.05 | |

| | Mean | 95% CI | *p-value* |
|---|---|---|---|
| **Total n-6/n-3** | | | |
| **Low LA** | 17.69 | 16.3, 19.1 | 0.73 |
| **High LA** | 18.04 | 16.6, 19.5 | |
| **ARA/EPA** | | | |
| **Low LA** | 15.39 | 13.6, 17.2 | 0.312 |
| **High LA** | 16.81 | 14.5, 19.1 | |
| **MUFA** | | | |
| **OA** | | | |
| **Low LA** | 45.8 | 42.2, 49.4 | 0.091 |
| **High LA** | 51.4 | 45.6, 57.3 | |
